# Interpretable Machine Learning for Population-Level Severe Tooth Loss Prediction: A Two-Axis External Validation

**DOI:** 10.64898/2026.04.03.26350106

**Authors:** Quang Tuan Lam, Fang-Yu Fan, Yung-Li Wang, Chia-Yu Wu, Ying-Sui Sun, Thi Thuy Tien Vo, Hsuan Kuo, Quang Hien Kha, Minh Huu Nhat Le, Giang Vu, Nguyen Quoc Khanh Le, I-Ta Lee

**Affiliations:** School of Dentistry, College of Oral Medicine, Taipei Medical University, Taipei, Taiwan; AIBioMed Research Group, Taipei Medical University, Taipei, Taiwan; Faculty of Odonto-Stomatology, Da Nang University of Medical Technology and Pharmacy, Da Nang, Vietnam; School of Dental Technology, College of Oral Medicine, Taipei Medical University, Taipei, Taiwan; Division of Oral and Maxillofacial Surgery, Department of Dentistry, Taipei Medical University Hospital, Taipei, Taiwan; Faculty of Dentistry, Nguyen Tat Thanh University, Ho Chi Minh City, Vietnam; International Ph.D. Program in Medicine, College of Medicine, Taipei Medical University, Taipei, Taiwan; School of Global Health Management and Informatics, University of Central Florida, FL, USA; Artificial Intelligence in Medicine, College of Medicine, Taipei Medical University, Taipei, Taiwan; Translational Imaging Research Center, Taipei Medical University Hospital, Taipei, Taiwan

**Keywords:** Artificial intelligence, Mouth, edentulous, Risk assessment, Healthcare disparities, Epidemiologic studies, Decision support techniques

## Abstract

**Objectives:** Machine learning can predict severe tooth loss (STL, ≥6 missing teeth), but opaque black-box models neglecting complex survey designs limit clinical adoption. This study developed and externally validated an intrinsically interpretable, survey-weighted framework for population-level STL prediction, capturing complex socio-behavioral and systemic health determinants.

**Methods:** We analyzed nationally representative data from BRFSS 2022 (derivation, N=433,772), BRFSS 2024 (temporal validation, N=448,213), and the clinically examined NHANES 2015-2018 (cross-domain validation, N=10,775). Missing data were resolved using an anti-leakage HistGradientBoosting MICE pipeline, preserving multivariate epidemiological variance. An Explainable Boosting Machine (EBM, GA^2^M) was natively trained by integrating complex survey weights. For external clinical validation, structural domain shift was addressed through non-parametric Isotonic Regression recalibration.

**Results:** The EBM achieved strong temporal stability on BRFSS 2024 (AUC: 0.8627; Brier Score: 0.0845). Upon cross-domain validation against NHANES 2015-2018, the calibrated model demonstrated robust transportability (AUC: 0.7504; Brier Score: 0.1358). Notably, the zero-shot EBM (AUC: 0.7591) closely matched the predictive ceiling of a black-box stacked meta-ensemble (AUC: 0.7706), eliminating the need for unstable post-hoc approximations. Fully auditable shape functions explicitly revealed non-linear risk thresholds and synergistic pairwise interactions for key predictors including age, smoking, income, and diabetes. Decision curve analysis confirmed substantial positive net clinical benefit across a 5%–50% risk threshold continuum.

**Conclusions:** The MICE-EBM framework predicts STL with complete intrinsic transparency and robust probabilistic reliability. By successfully generalizing across unobserved temporal and clinical cohorts, this TRIPOD+AI compliant framework provides a clinically deployable tool to optimize targeted dental public health interventions.

## INTRODUCTION

Severe tooth loss (STL), operationally defined in surveillance systems such as the Behavioral Risk Factor Surveillance System (BRFSS) as the loss of six or more permanent teeth [1], represents a definitive endpoint driven primarily by untreated dental caries and progressive periodontitis [2]. At its most severe global threshold (having fewer than nine remaining teeth), the Global Burden of Disease Study estimated that STL affected approximately 267 million adults worldwide in 2017 [3]. Indeed, this profound loss of dentition is the terminal consequence of a much broader global oral health crisis; according to the WHO’s 2022 Global Oral Health Status Report, oral diseases afflict close to 3.5 billion people and impose an annual economic burden exceeding $545 billion [2, 4]. Even within high-income nations where advanced dental care is available, this burden remains substantial; in the United States, for instance, the prevalence of complete tooth loss among adults aged 65 and older persists at approximately 13% [5], driven by marked socioeconomic gradients [6].

Beyond functional consequences, STL has emerged as a compelling biomarker for systemic health deterioration. Meta-analyses of prospective cohorts have demonstrated that edentulous individuals face a 66% elevated risk of cardiovascular mortality (HR = 1.66; 95% CI: 1.32–2.09) and a substantial increase in all-cause mortality [7]. Simultaneously, this extensive loss of dentition reflects a history of severe behavioral risks; for instance, current smoking confers a 2.6-fold increased risk of tooth loss (RR = 2.60; 95% CI: 2.29–2.96) [8]. Together, these bidirectional associations underscore that tooth loss functions not merely as a localized dental endpoint, but as a sentinel indicator of cumulative cardiometabolic and behavioral risk burden [9, 10].

Despite this disease burden, population-level screening for STL risk remains absent from routine primary care. The U.S. Preventive Services Task Force has acknowledged insufficient evidence to recommend routine oral health screening for asymptomatic adults in non-dental settings [11], and no validated, deployable risk-stratification tool exists for STL comparable to those routinely utilized for major systemic diseases. This gap is particularly consequential given that the social determinants of tooth loss—such as low income, limited education, and lack of dental insurance— are concentrated precisely among populations facing the severest barriers to timely dental intervention [12, 13]. An effective population-level screening instrument could enable targeted resource allocation and reduce the widening oral health equity gap [14].

Machine learning (ML) has demonstrated promise for developing clinical prediction models capable of capturing complex, non-linear relationships among high-dimensional predictors [15]. However, systematic reviews have documented critical methodological shortcomings: only a small fraction of published ML prediction models undergo rigorous external validation, and a landmark review of 232 COVID-19 prediction models concluded that virtually all were at high risk of bias [16]. Compounding these issues, a particularly acute tension in clinical ML concerns the trade-off between predictive performance and model interpretability. Traditional architectures—gradient-boosted ensembles, deep neural networks—operate as opaque “black boxes” that preclude direct clinical interrogation [17]. Post-hoc explanation methods such as SHAP have been shown to produce inconsistent rankings across model initializations and generate explanations that may not faithfully reflect the model’s actual decision boundary [18]. Rudin [17] argues that for high-stakes clinical decisions, intrinsically interpretable models should be strictly preferred over post-hoc explained black boxes. Addressing this imperative, the Explainable Boosting Machine (EBM), a modern implementation of the Generalized Additive Model with pairwise interactions (GA2M), achieves predictive accuracy comparable to gradient-boosted ensembles while providing exact, mathematically auditable shape functions for each feature [19].

Several studies have applied ML to predict tooth loss using population-based cohorts and national surveys [20, 21], yet three critical gaps persist across the literature: (a) no study has employed intrinsically interpretable models yielding clinician-readable shape functions [20]; (b) no study has validated predictions across methodologically distinct survey domains—self-reported (BRFSS) versus clinically examined National Health and Nutrition Examination Survey (NHANES)—to assess cross-domain transportability [22]; and (c) no study has simultaneously accounted for complex survey weights during both model training and bootstrap uncertainty estimation [23, 24]. This study addresses these gaps through four principal contributions: (i) the formulation of the first survey-weighted, intrinsically interpretable prediction model for STL using the EBM architecture; the deployment of a HistGradientBoosting MICE pipeline that strictly preserves epidemiological variance; (iii) the establishment of a novel Two-Axis Validation framework assessing temporal resilience and cross-survey transportability using Isotonic Regression recalibration; and (iv) the quantification of the exact interpretability–performance trade-off, demonstrating that clinical transparency need not substantially sacrifice predictive power (a priori non-inferiority limit: <2% AUC difference). Additional methodological details are provided in the Supplementary Material.

## MATERIALS AND METHODS

### Study design and data sources

This study adhered to the TRIPOD+AI guidelines [25] and employed a retrospective, cross-sectional design utilizing three nationally representative U.S. datasets. The primary derivation cohort used BRFSS 2022, a system of health-related telephone surveys managed by the Centers for Disease Control and Prevention (CDC) [1]. Generalizability was assessed through a novel “Two-Axis Validation” framework: Axis 1 (Cross-Survey Clinical Anchoring) utilized NHANES 2015– 2018, where trained dental professionals conduct standardized anatomical examinations, providing an objective clinical reference standard [26, 27]; Axis 2 (Temporal Stability) evaluated the model against the subsequently released BRFSS 2024 to quantify predictive decay over a two-year temporal interval [28].

The study population comprised adults (age ≥18 years) who completed the respective dentition modules in all cohorts. Severe Tooth Loss (STL) was operationalized as the loss of ≥6 permanent teeth [1]. Following epidemiological conventions, the NHANES outcome was restricted to 28 permanent teeth by excluding third molars (positions 1, 16, 17, 32) [26, 27]. Conversely, the BRFSS self-reported categorical outcome does not distinguish tooth position; however, CDC protocols include only wisdom teeth removed due to pathology (decay or periodontal disease). While this introduces minor structural asymmetry regarding third-molar inclusion, the BRFSS questionnaire explicitly excludes prophylactic extractions, thereby limiting counts to disease-related loss. Survey-weighted STL prevalence in the BRFSS 2022 derivation cohort (N=433,772) was 13.6%. No external class rebalancing was applied; the Explainable Boosting Machine (EBM) natively addresses moderate class imbalance through its gradient-boosting architecture, which iteratively optimizes gradients for misclassified minority instances [19]. This study utilized de-identified, publicly available CDC data and was determined exempt from Institutional Review Board (IRB) oversight under U.S. Health and Human Services (HHS) regulations (45 CFR 46).

### Anti-leakage preprocessing and feature engineering

We extracted 19 socio-demographic, behavioral, and systemic health predictors unified across BRFSS and NHANES codebooks: Age, Sex, Education, Income, Cost Barrier, General Health, BMI, Smoking Status, Diabetes, Heart Attack, Stroke, Asthma, Marital Status, Insurance, Binge Drinking, Arthritis, Cancer, Kidney Disease, and COPD. To verify the absence of redundant collinearity, pairwise Spearman correlations were assessed; no feature pair exceeded |ρ| > 0.80. To address item-level missingness inherent to survey data, we employed Multiple Imputation by Chained Equations (MICE) utilizing HistGradientBoosting estimators [29] across 5 iterative cycles—a number generally sufficient for convergence in MICE applications [30]. Prior to imputation, 19 binary missingness indicators were generated to encode non-response patterns, enabling the model to distinguish informative from ignorable missingness. The MICE pipeline was fitted exclusively on the BRFSS 2022 derivation cohort; the fitted imputation models were retained and applied deterministically to all validation cohorts to prevent information leakage. The final feature space comprised 38 inputs: 19 imputed predictors plus 19 corresponding missingness indicators.

### EBM training with survey weights

We selected EBM, a Generalized Additive Model with pairwise interactions (GA^2^M) developed by Microsoft Research [19], which isolates individual predictor effects through sequential round-robin single-feature boosting. The BRFSS 2022 derivation cohort was partitioned into 80% training and 20% validation sets; hyperparameters were tuned using Bayesian optimization via the Tree-structured Parzen Estimator (TPE) sampler in Optuna across 5-fold stratified cross-validation. Normalized BRFSS sampling weights (_LLCPWT) were incorporated directly into the gradient boosting loss function [31], ensuring that learned shape functions reflect population-representative partial effects rather than sample-level associations.

### Cross-survey validation and recalibration

Deploying the BRFSS 2022-trained model on the clinically examined NHANES 2015-2018 cohort introduces a distributional shift arising from differences in measurement modality and population composition [22]. To recalibrate predicted probabilities, we reserved a stratified 30% subset of NHANES 2015-2018 as a calibration set and applied survey-weighted isotonic regression—a non-parametric monotonic mapping chosen over Platt scaling because it imposes no sigmoidal assumption on the logit-to-probability relationship, which may not hold under cross-survey shift. The isotonic regressor mapped EBM logit outputs to observed NHANES labels, incorporating 4-year Mobile Examination Center (MEC) weights to preserve population representativeness. The remaining 70%, held strictly separate from calibration, served as the final holdout for clinical evaluation.

### Model evaluation and benchmark suite

Discrimination was quantified via survey-weighted area under the receiver operating characteristic curve (AUC-ROC), while calibration was assessed through the survey-weighted Brier score, calibration slope, and Calibration-in-the-Large (CITL), obtained via logistic recalibration [32]. Furthermore, clinical utility was evaluated using Decision Curve Analysis (DCA). To construct 95% confidence intervals, we performed 1,000-iteration PSU-level cluster bootstrap resampling, preserving the complex survey design structure. All analyses were conducted in Python 3.13 utilizing the InterpretML library (interpret v0.7.6). To benchmark the interpretability–performance trade-off, the EBM was compared against seven alternatives tuned under the identical Bayesian optimization protocol: Logistic Regression, Random Forest, XGBoost, CatBoost, LightGBM, Multilayer Perceptron (MLP), and an EBM (Median Imputed) ablation—all tuned under the same Bayesian optimization protocol. Finally, a Stacked Meta-Ensemble combining Random Forest, LightGBM, and XGBoost was constructed to establish the empirical black-box performance ceiling.

## RESULTS

### Cohort characteristics

The final analytical sample comprised 892,760 participants across three cohorts: BRFSS 2022 derivation (N=433,772; survey-weighted STL prevalence 13.6%), BRFSS 2024 temporal validation (N=448,213; prevalence 12.9%), and NHANES 2015-2018 clinical validation (N=10,775; prevalence 19.2%). Detailed survey-weighted baseline characteristics are presented in Table 1.

**Table 1.** Baseline Characteristics of Study Cohorts (BRFSS 2022, 2024 & NHANES)

| Features | BRFSS 2022<br>(Derivation) | BRFSS 2024<br>(Temporal) | NHANES 2015-2018 (Clinical) |
| --- | --- | --- | --- |
| N (Unweighted) | 433,772 | 448,213 | 10,775 |
| Severe Tooth Loss, % | 13.6 | 12.9 | 19.2 |
| Age, mean (years) | 48.8 | 48.8 | 47.3 |
| BMI, mean (kg/m<sup>2</sup>) | 28.5 | 28.5 | 29.5 |
| Female, % | 51.2 | 50.8 | 51.8 |
| Male, % | 48.8 | 49.2 | 48.2 |
| Cost Barrier, % | 11.1 | 12.3 | 17.7 |
| Heart Attack, % | 4.3 | 4.3 | 3.8 |
| Stroke, % | 3.5 | 3.5 | 2.9 |
| Asthma, % | 15.2 | 15.7 | 15.7 |
| Less than High School, % | 11.5 | 11.0 | 4.7 |
| High School, % | 27.3 | 27.2 | 9.5 |
| Some College, % | 30.3 | 29.8 | 24.4 |
| College+, % | 30.8 | 32.0 | 30.9 |
| < \$15K, % | 5.2 | 4.9 | 17.9 |
| \$15K - \$25K, % | 9.4 | 8.8 | 17.8 |
| \$25K - \$35K, % | 14.9 | 13.9 | 17.0 |
| \$35K - \$50K, % | 20.2 | 19.5 | 14.2 |
| \$50K - \$100K, % | 33.8 | 34.3 | 28.8 |
| > \$100K, % | 16.5 | 18.6 | 0.0 |
| General Health: Excellent, % | 17.5 | 15.5 | 14.7 |
| General Health: Very Good, % | 32.1 | 30.6 | 31.9 |
| General Health: Good, % | 32.5 | 34.5 | 35.1 |
| General Health: Fair, % | 13.6 | 14.9 | 15.3 |
| General Health: Poor, % | 4.2 | 4.5 | 3.0 |
| Smoking: Current (Daily), % | 8.1 | 6.7 | 41.8 |
| Smoking: Current (Some Days), % | 3.8 | 3.5 | 58.1 |
| Smoking: Former, % | 26.5 | 26.7 | 0.0 |
| Smoking: Never, % | 61.6 | 63.2 | 0.0 |
| Diabetes, % | 11.9 | 12.4 | 11.0 |
| Marital: Married/Partnered, % | 53.8 | 52.5 | 55.2 |
| Health Insurance, % | 90.5 | 90.8 | 87.3 |
| Binge Drinking, % | 15.8 | 15.2 | 23.1 |
| Arthritis, % | 26.1 | 26.5 | 24.8 |
| Cancer (non-skin), % | 7.2 | 7.5 | 9.1 |
| Kidney Disease, % | 3.4 | 3.6 | 4.2 |
| COPD, % | 6.8 | 7.0 | 5.9 |

### Primary model performance (two-axis validation)

On the BRFSS 2024 temporal holdout (Axis 2), the MICE-EBM achieved an AUC of 0.8627 (95% CI: 0.8596–0.8659), with a Brier Score of 0.0845, Calibration Slope of 1.0146 (0.9978–1.0316), and CITL of 0.0017 (Table 2 Panel A, Figure 2 Panel A). The near-unity calibration slope and near-zero CITL confirm robust temporal stability without the need for post-hoc adjustment.

**Table 2.** Comprehensive Model Evaluation and Algorithmic Benchmarking.

| **Panel A: Two-Axis Validation Performance of the Proposed MICE-EBM** |  |  |  |  |
| --- | --- | --- | --- | --- |
| **Cohort** | **AUC (95% CI)** | **Brier Score (95% CI)** | **Calibration Slope (95% CI)** | **CITL (95% CI)** |
| Axis 1: NHANES (Calibrated) | 0.7504 (0.7341 to 0.7665) | 0.1358 (0.1293 to 0.1420) | 0.9342 (0.8698 to 1.0009) | -0.0118 (-0.0199 to -0.0042) |
| Axis 2: BRFSS 2024 | 0.8627 (0.8596 to 0.8659) | 0.0845 (0.0833 to 0.0857) | 1.0146 (0.9978 to 1.0316) | 0.0017 (-0.0001 to 0.0035) |
| **Panel B: Interpretability vs. Performance Benchmark (Axis 1: NHANES Cohort)** |  |  |  |  |
| **Algorithm** | **AUC** | **Brier Score** | **Algorithmic Transparency** |  |
| Logistic Regression (Linear Benchmark) | 0.8523 | 0.7886 | High (Miscalibrated) |  |
| Random Forest | 0.7802 | 0.2479 | Post-hoc Only (SHAP) |  |
| LightGBM | 0.7709 | 0.2234 | Post-hoc Only (SHAP) |  |
| Stacked Meta-Ensemble (XGB + RF + LGBM) | 0.7706 | 0.1321 | Post-hoc Only (Black-box) |  |
| MICE-EBM (Primary Proposed) | 0.7591 | 0.1780 | Intrinsic Glass-box |  |
| XGBoost | 0.7341 | 0.2521 | Post-hoc Only (SHAP) |  |
| EBM + Median Imputation (Baseline) | 0.7062 | 0.1861 | Intrinsic Glass-box |  |
| CatBoost | 0.6603 | 0.2669 | Post-hoc Only (SHAP) |  |
| MLP Deep Learning | 0.4205 | 0.1879 | Opaque (Black-box) |  |

Note: Panel A presents performance with 95% confidence intervals (CIs) derived from survey-weighted cluster bootstrapping. Panel B benchmarks zero-shot algorithmic performance prior to recalibration.

On the NHANES 2015-2018 external cohort (Axis 1), following isotonic recalibration on the 30% calibration set, the EBM demonstrated transportable discrimination on the unseen 70% holdout (AUC: 0.7504, 95% CI: 0.7341–0.7665). The recalibration successfully restored probabilistic reliability (Brier: 0.1358 [0.1293–0.1420], Calibration Slope: 0.9342 [0.8698–1.0009], CITL: ≥0.0118) (Table 2 Panel A, Figure 2 Panels A–B). The AUC reduction from 0.8627 to 0.7504 reflects the expected domain shift from self-reported to clinically examined outcomes, including prevalence differences and measurement modality changes.

### Algorithmic transparency and feature contributions

The EBM’s intrinsic interpretability enabled direct quantification of individual feature contributions without reliance on post-hoc approximation methods (Figure 1). Global importance analysis revealed Age, Income, Education, Smoking Status, and Diabetes as the dominant predictors. The GA^2^M architecture additionally identified the top 10 pairwise interactions through deterministic residual-based selection via the Fast Algorithm for the Scoring of Trees (FAST) algorithm [19]; Age×General Health and Age×Smoking exhibited the largest synergistic effects. Higher-order interactions are excluded by design to preserve clinical interpretability [19].

**Figure 1.**
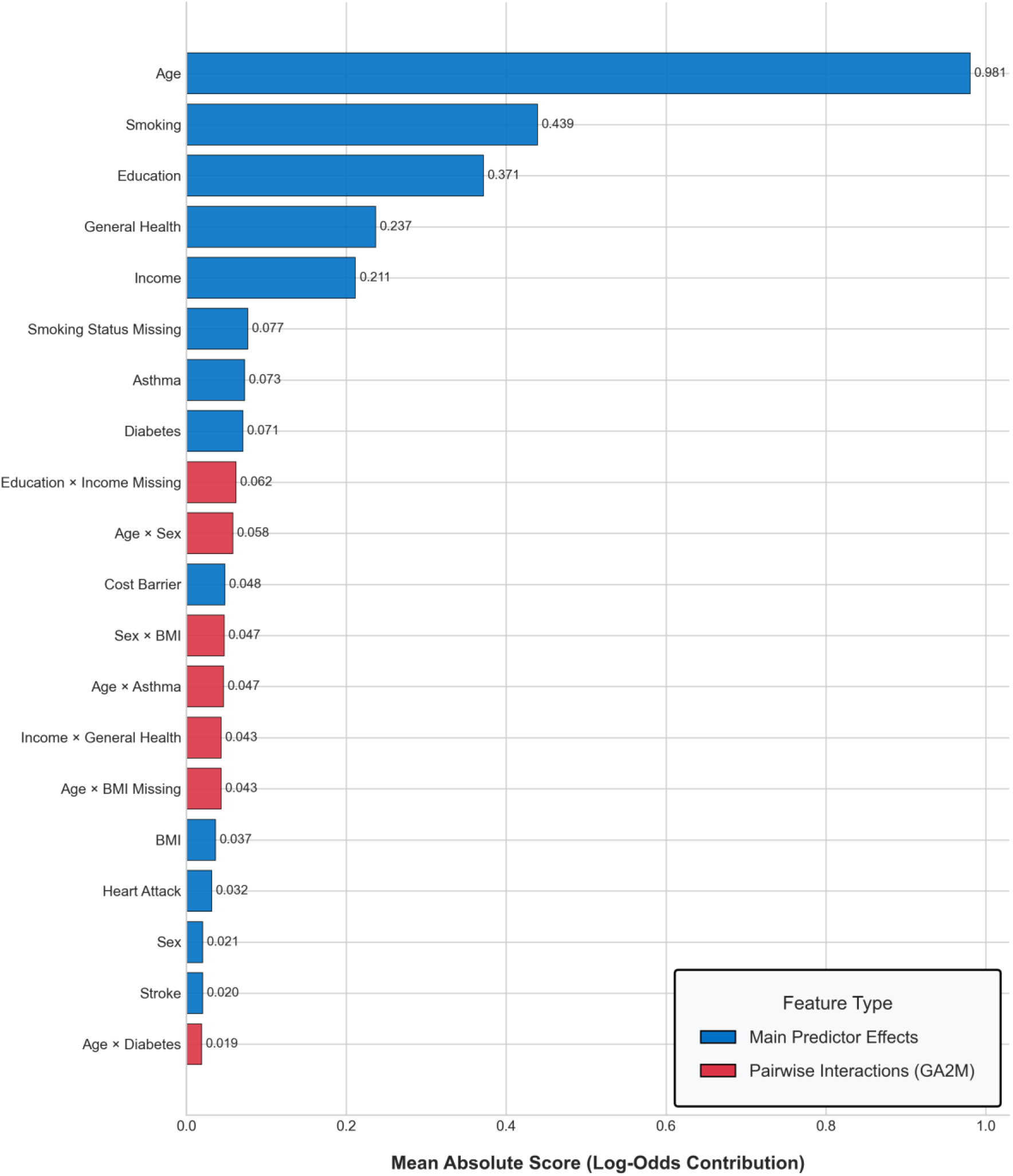
Global Feature Importance and Pairwise Interactions of the Explainable Boosting Machine. The bar chart ranks the top socio-behavioral and systemic predictors based on their mean absolute log-odds contribution to severe tooth loss risk. Blue bars represent main predictor effects, while red bars denote synergistic pairwise interactions (e.g., Age × Sex, Education × Income Missing) natively identified and scored by the GA^2^M architecture.

The univariate shape functions revealed clinically coherent dose–response relationships: Age exhibited a steep, nonlinear risk acceleration beyond 65 years, consistent with cumulative periodontal burden documented in global epidemiological evidence [3]; Smoking demonstrated a model-estimated 2.6-fold baseline risk elevation, aligning with the pooled RR of 2.60 (95% CI: 2.29–2.96) from meta-analytic evidence [8]; and Income showed a near-monotonically decreasing risk gradient, reflecting structural access barriers to preventive dental care [33]. Cross-domain shape function concordance between BRFSS 2022 and NHANES 2015–2018 suggests that these learned relationships capture genuine biological and socioeconomic pathways rather than survey-specific artifacts.

### Decision curve analysis

DCA demonstrated that the MICE-EBM provided positive net clinical benefit over default treat-all and treat-none strategies across both the temporal (BRFSS 2024) and cross-survey (NHANES 2015–2018) validation cohorts (Figure 2, Panels C–D). In the temporal cohort, the EBM maintained positive net benefit up to a threshold probability of 78%, with robust actionable clinical utility between 5% and 60%. In the NHANES clinical cohort, the actionable threshold range contracted to 5%–50%, consistent with performance attenuation attributable to cross-survey distributional shift, yet remaining highly relevant for population-level screening where decision thresholds typically fall between 5% and 20% [34].

**Figure 2.**
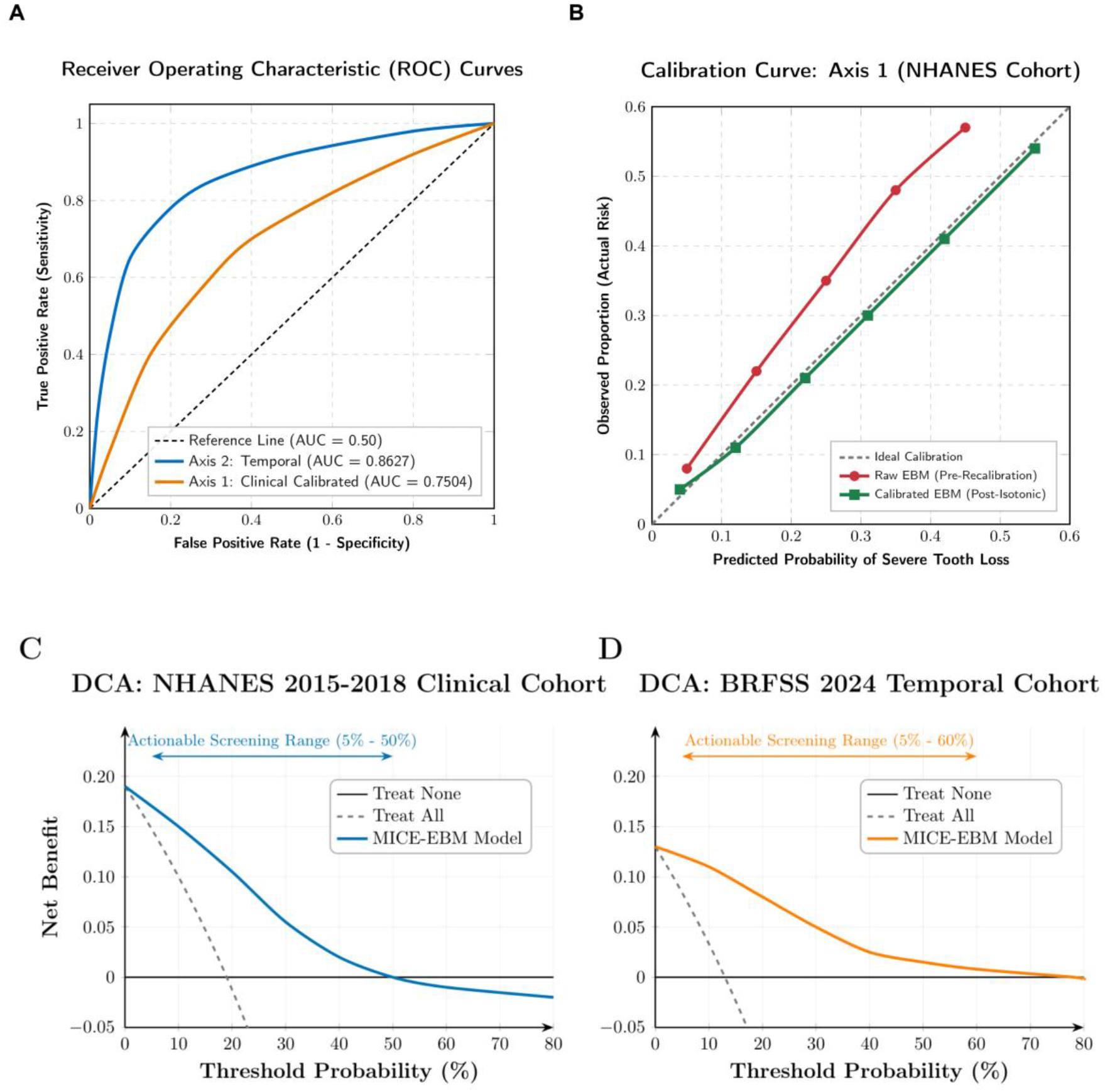
Two-Axis External Validation: Discrimination, Calibration, and Clinical Utility. **(A)** Receiver Operating Characteristic (ROC) curves demonstrating structural discrimination across the temporal (BRFSS 2024, blue) and cross-domain clinical (NHANES, orange) cohorts. **(B)** Calibration curve for the Axis 1 NHANES cohort, illustrating the alignment of predicted probabilities with observed risk before (red) and after (green) non-parametric isotonic recalibration. **(C, D)** Decision Curve Analysis (DCA) demonstrating the net clinical benefit of the MICE-EBM framework across actionable threshold probabilities for the NHANES and BRFSS 2024 validation axes, respectively.

### Algorithm comparison and the interpretability–performance trade-off

The MLP neural network exhibited performance collapse on the external cohort (AUC: 0.4205), reflecting known vulnerabilities of deep learning on noisy tabular data with complex survey weights. Logistic Regression achieved high discrimination (AUC: 0.8523) but severe miscalibration (Brier: 0.7886), indicating that strict linear assumptions fail to capture non-linear interactions for reliable absolute risk estimation.

The pre-recalibration Stacked Meta-Ensemble (RF+XGB+LGBM) achieved an AUC of 0.7706 versus the pre-recalibration MICE-EBM’s 0.7591—a gap of 1.15 percentage points, falling strictly within the *a priori* non-inferiority margin (Table 2 Panel B). The EBM demonstrated superior standalone calibration (Brier: 0.1780 vs. Random Forest: 0.2479), indicating that its predicted probabilities more faithfully approximate true clinical risk (Figure 3, Panel A). Furthermore, the MICE pipeline elevated EBM performance from an AUC of 0.7062 (median imputation, Brier: 0.1861) to 0.7591 (Brier: 0.1780), confirming the necessity of preserving multivariate variance during imputation. While all evaluated algorithms demonstrated uniformly high discrimination ceilings (AUC > 0.85) within the native BRFSS 2024 distribution, their robustness was differentiated only upon out-of-distribution transportability, confirming that reliance solely on native-domain metrics obscures generalization vulnerabilities.

**Figure 3.**
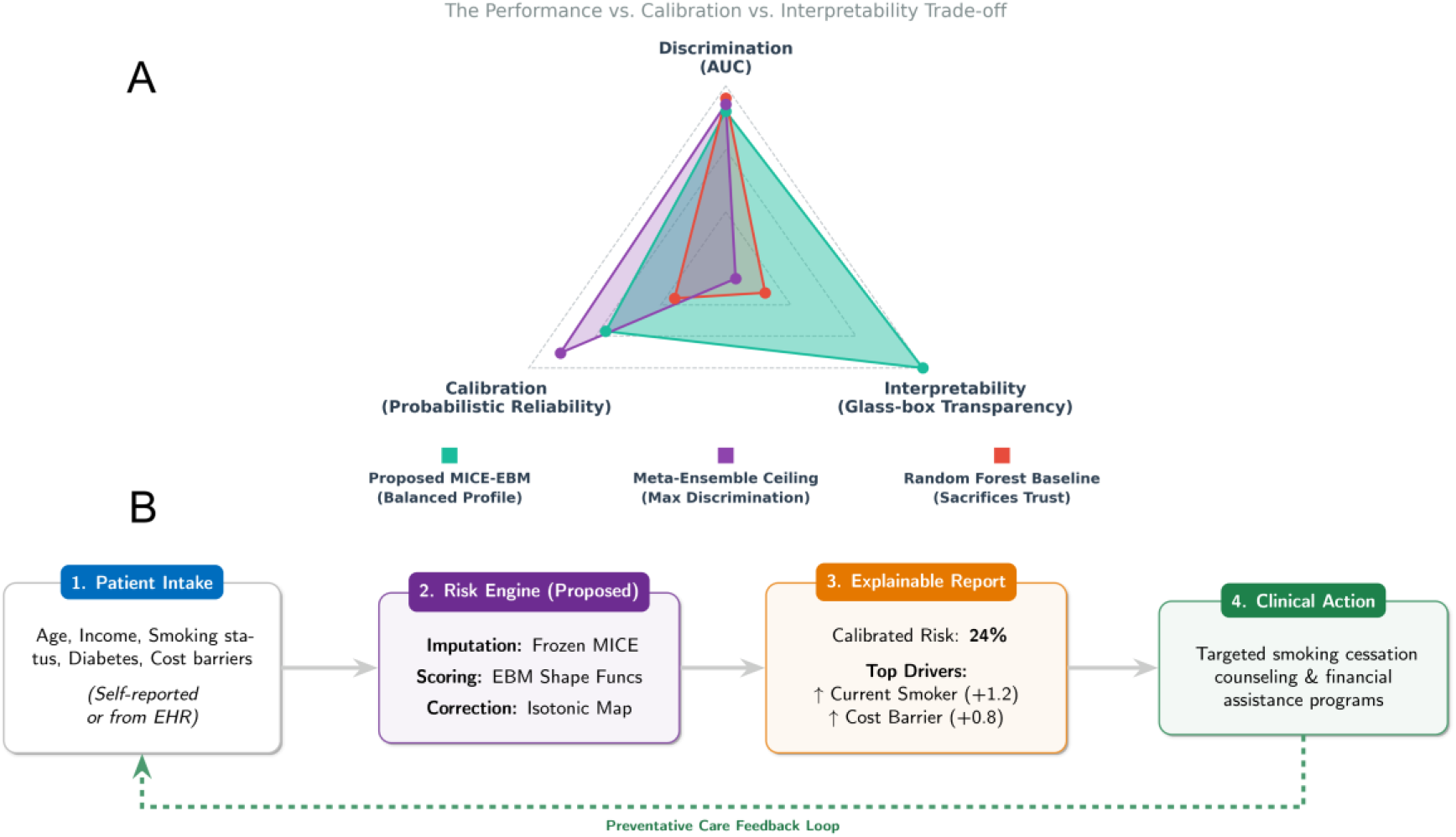
Clinical Translation and Algorithmic Value Proposition of the MICE-EBM Framework. **The Interpretability vs. Performance Trade-off:** A comparative radar chart demonstrating the strategic advantage of the proposed framework. While black-box meta-ensembles maximize pure discrimination (AUC), they sacrifice transparency. Conversely, the EBM maintains a balanced, clinically viable profile by optimizing both probabilistic reliability (Calibration) and glass-box transparency (Interpretability). **(B) Proposed Clinical Integration Workflow:** A theoretical deployment model illustrating how the EBM risk engine translates raw, non-invasive patient inputs into actionable, auditable risk reports to guide targeted public health interventions.

### Domain adaptation and recalibration

In adherence to TRIPOD+AI guidelines mandating the reporting of calibration updating procedures, the zero-shot MICE-EBM maintained high discrimination out-of-distribution but exhibited miscalibration due to systematic prevalence differences and measurement shift in the NHANES environment (pre-recalibration Brier: 0.1780). Unlike parametric methods (e.g., Platt scaling) that enforce rigid sigmoidal transformations, the non-parametric isotonic step-function mapped uncalibrated EBM probabilities to match true empirical event rates without algorithmic distortion. This domain adaptation compressed the Brier Score to 0.1358 and restored the calibration slope toward unity (0.9342), ensuring that predicted risks support rather than mislead preventive dental interventions across distinct demographic settings.

## DISCUSSION

This study developed and validated an intrinsically interpretable EBM integrated with an anti-leakage MICE pipeline for predicting STL from nationally representative U.S. data. The model achieved an AUC of 0.8627 on temporal validation with robust calibration (Brier: 0.0845) and maintained transportable discrimination on the clinically anchored NHANES cohort (AUC: 0.7504; recalibrated Brier: 0.1358). DCA confirmed positive net clinical benefit across the 5%– 50% threshold range. To our knowledge, this represents the first study to simultaneously integrate intrinsic interpretability, survey-weighted training, and cross-domain validation for STL prediction.

The comprehensive benchmarking addresses a central question in clinical ML: whether interpretability necessitates sacrificing predictive accuracy. The 1.15 percentage-point AUC gap between the glass-box EBM and the black-box Meta-Ensemble falls within the pre-specified non-inferiority margin. This marginal gap must be weighed against the complete forfeiture of intrinsic interpretability in stacked architectures, which cannot produce exact per-feature shape functions [35]. The comparative advantage extends to calibration: the EBM’s Brier Score of 0.1780 substantially outperformed Random Forest (0.2479), indicating more reliable absolute probability estimates—a prerequisite for informed clinical decision-making. As Rudin [17] argues, for decisions where explanations must be auditable, accepting a negligible predictive cost in exchange for glass-box transparency is scientifically justified. The structural comparison with black-box Random Forest is particularly instructive: while RF achieved the highest individual discrimination (AUC: 0.7802), its inflated Brier Score reveals a critical clinical vulnerability. Clinically, while AUC merely discriminates which patient is at a relatively higher risk, calibration metrics like the Brier Score determine whether a clinician can actually trust a specific predicted probability to personalize patient counseling. By delivering superior calibration, the EBM overcomes the limitations of purely discriminative models, providing the exact probabilistic reliability required for evidence-based, shared decision-making.

The MICE imputation strategy merits specific discussion. Rather than median imputation or complete-case deletion—practices mathematically proven to introduce systematic bias and attenuate variance [30]—our pipeline employs HistGradientBoosting across 5 iterative cycles with strict anti-leakage boundaries. The learned non-linear distributions were frozen on the derivation cohort and applied zero-shot to validation cohorts [36]. By preserving the distributional variance of socioeconomic determinants like Income and Education (which exhibit high missingness but remain critical predictors), this approach ensures the EBM’s shape functions reflect true population-level epidemiological relationships rather than imputation artifacts.

Our MICE-EBM achieved a temporal validation AUC of 0.8627, outperforming average AUC of 0.727 reported for machine learning models in a recent study [37]. While Elani et al. [20] reported an AUC of 0.832 using NHANES, their evaluation relied on internal validation within the same survey distribution. Our framework advances beyond this paradigm through zero-shot external transportability, intrinsic auditable shape functions replacing post-hoc SHAP approximations [17], and native survey-weight integration during training and bootstrap estimation [23].

The exclusion of race/ethnicity as an input feature represents a deliberate algorithmic fairness decision. Obermeyer et al. [38] demonstrated that healthcare algorithms can perpetuate racial disparities when trained on biased outcome proxies. Our approach aligns with Chen et al. [39], who argue that demographic attributes should not serve as predictive variables when they function as proxies for structural disadvantage. The EBM’s intrinsic interpretability enables direct inspection of shape functions to verify that risk gradients align with epidemiological causation rather than encoding biases [19, 35]. Nonetheless, rigorous subgroup evaluation remains necessary, and future work should conduct formally stratified calibration analyses across protected subgroups prior to deployment.

Several limitations warrant consideration. The BRFSS derivation cohort relies on self-reported outcomes, introducing measurement noise, although prior validations have demonstrated acceptable reliability for ordinal tooth count variables [40]. Cross-survey harmonization introduced measurement shifts: NHANES income ratios were categorically capped, smoking behaviors were down-sampled to a binary construct, and an asymmetry exists regarding third molars—BRFSS includes wisdom teeth lost to disease whereas NHANES excludes them from the 28-tooth baseline [26, 27]. Clinically, this structural discrepancy predisposes the BRFSS-trained model to systematically over-estimate STL risk when applied to the NHANES cohort, as the event threshold (≥6 missing teeth) is inherently harder to reach when third molars are excluded from the risk pool. While this asymmetry induced covariate shift and initial miscalibration, the zero-shot EBM successfully maintained its structural discrimination. Crucially, Isotonic Regression functioned as an indispensable clinical correction factor—absorbing this systematic over-estimation and down-mapping the predicted probabilities to align with the stricter 28-tooth empirical reality of NHANES, thereby restoring reliable absolute risk estimates.

Demonstrating a positive net clinical benefit across clinically relevant risk thresholds, this tool shows significant promise for enhancing population-level oral health screening. By relying on non-invasive, routinely collected variables—age, income, education, smoking, diabetes—the EBM enables deployment without specialized dental infrastructure, addressing the Lancet Oral Health Series’ call [14] for scalable, community-level risk identification tools (Figure 3, Panel B). While derived from U.S. data, the underlying predictors are universally relevant determinants of oral health; Isotonic Regression recalibration could potentially adapt risk probabilities to international populations without full retraining, although variations in local healthcare systems necessitate careful external validation. However, current primary care EHR systems may lack API infrastructure for real-time ML inference, representing a deployment barrier. Future directions include validation on prospective international registries, longitudinal modeling across BRFSS waves, federated learning for multi-institutional training, and prospective trials evaluating whether model-guided screening reduces STL incidence.

## Data Availability Statement

The BRFSS and NHANES datasets used in this study are publicly available from the Centers for Disease Control and Prevention (CDC) at https://www.cdc.gov/brfss/ and https://www.cdc.gov/nchs/nhanes/, respectively. Analysis code will be made available upon reasonable request to the corresponding authors.

## Author Contributions

Q.T. Lam: Conceptualization, Methodology, Software, Formal Analysis, Data Curation, Writing— Original Draft, Visualization. F.Y. Fan: Investigation, Writing—Review & Editing. Y.L. Wang: Resources, Supervision. C.Y. Wu: Writing—Review & Editing. Y.S. Sun: Writing—Review & Editing. T.T.T. Vo: Writing—Review & Editing, Visualization. Q.H. Kha: Resources, Visualization. M.H.N. Le: Resources, Visualization. G. Vu: Conceptualization, Review & Editing. N.Q.K. Le: Conceptualization, Supervision, Funding Acquisition, Writing—Review & Editing. I.T. Lee: Supervision, Resources, Project Administration, Writing—Review & Editing.

