## Supplemental Table S1 to S3; Supplemental Figure S1 to S8 for "Interpretable Machine Learning for Population-Level Severe Tooth Loss Prediction: A Two-Axis External Validation"

**SUPPLEMENTARY MATERIAL**

***PART A: SUPPLEMENTARY TABLES***

*Table S1: TRIPOD+AI Reporting Compliance Checklist*

| TRIPOD Item | Description | Status |
| --- | --- | --- |
| **1** | Title: Prediction model type | ✓ |
| **2** | Abstract: Structured summary | ✓ |
| **3a** | Objective: Study objectives | ✓ |
| **4a** | Source of data | ✓ BRFSS/NHANES |
| **5a** | Participants: Setting/dates | ✓ |
| **6a** | Outcome: Definition | ✓ ≥ 6 teeth |
| **7a** | Predictors: Definition | ✓ 19 features |
| **8** | Sample size | ✓ N = 892,760 |
| **9** | Missing data handling | ✓ MICE (m=5) |
| **10a** | Model specification | ✓ EBM (GA2M) |
| **10b** | Model training | ✓ Survey-weighted |
| **12** | Validation strategy | ✓ Two-Axis |
| **14a** | Discrimination | ✓ AUC + 95% CI |
| **14b** | Calibration | ✓ Slope, CITL, Brier |
| **15a** | Model updating | ✓ Isotonic Regression |
| **16** | Limitations | ✓ Section 4.4 |
| **+AI-1** | Explainability | ✓ Intrinsic EBM |
| **+AI-2** | Fairness | ✓ Section 4.5b |

*Table S2: Predictor Spearman Correlation Matrix*

The following table presents the pairwise Spearman correlation coefficients (ρ) for all 19 core predictors evaluated in our study. This assessment ensures the absence of severe multicollinearity (|ρ| > 0.80) which could fragment feature importance attribution in Explainable Boosting Machines.

| Feature | Age | Sex | Marit | Educa | Incom | Insur | CostB | GenH | BMI | Smoke | Binge | Diabe | Heart | Strok | Asthm | Arthr | Cance | Kidn | COPD |
| --- | --- | --- | --- | --- | --- | --- | --- | --- | --- | --- | --- | --- | --- | --- | --- | --- | --- | --- | --- |
| Age | 1.00 | 0.06 | -0.20 | 0.02 | -0.02 | -0.13 | 0.15 | 0.13 | -0.01 | -0.07 | -0.15 | -0.16 | -0.16 | -0.13 | 0.05 | -0.37 | -0.24 | -0.12 | -0.14 |
| Sex | 0.06 | 1.00 | 0.03 | 0.02 | -0.03 | -0.04 | -0.02 | 0.02 | -0.05 | 0.07 | -0.07 | 0.00 | 0.06 | 0.00 | -0.07 | -0.10 | -0.03 | -0.01 | -0.03 |
| Marital | -0.20 | 0.03 | 1.00 | -0.15 | -0.18 | 0.14 | -0.10 | 0.09 | -0.05 | -0.03 | 0.05 | 0.02 | 0.01 | -0.01 | -0.05 | 0.06 | 0.05 | 0.00 | -0.02 |
| Education | 0.02 | 0.02 | -0.15 | 1.00 | 0.22 | -0.16 | 0.10 | -0.22 | -0.08 | 0.15 | -0.02 | 0.07 | 0.05 | 0.05 | 0.02 | 0.06 | -0.02 | 0.03 | 0.11 |
| Income | -0.02 | -0.03 | -0.18 | 0.22 | 1.00 | -0.00 | 0.12 | -0.18 | -0.08 | 0.21 | 0.13 | 0.07 | 0.06 | 0.06 | 0.05 | 0.10 | 0.02 | 0.05 | 0.11 |
| Insurance | -0.13 | -0.04 | 0.14 | -0.16 | -0.00 | 1.00 | -0.19 | 0.03 | -0.03 | 0.02 | 0.06 | 0.03 | 0.03 | 0.02 | 0.02 | 0.09 | 0.06 | 0.02 | 0.02 |
| Cost Barrier | 0.15 | -0.02 | -0.10 | 0.10 | 0.12 | -0.19 | 1.00 | -0.14 | -0.02 | 0.05 | -0.04 | -0.00 | 0.00 | 0.01 | 0.06 | -0.00 | -0.04 | 0.00 | 0.04 |
| Gen. Health | 0.13 | 0.02 | 0.09 | -0.22 | -0.18 | 0.03 | -0.14 | 1.00 | 0.23 | -0.14 | -0.04 | -0.21 | -0.15 | -0.14 | -0.13 | -0.25 | -0.11 | -0.14 | -0.22 |
| BMI | -0.01 | -0.05 | -0.05 | -0.08 | -0.08 | -0.03 | -0.02 | 0.23 | 1.00 | -0.02 | -0.02 | -0.16 | -0.03 | -0.02 | -0.09 | -0.13 | 0.00 | -0.05 | -0.04 |
| Smoking | -0.07 | 0.07 | -0.03 | 0.15 | 0.21 | 0.02 | 0.05 | -0.14 | -0.02 | 1.00 | 0.21 | 0.04 | 0.08 | 0.05 | 0.03 | 0.11 | 0.05 | 0.03 | 0.18 |
| Binge Drink | -0.15 | -0.07 | 0.05 | -0.02 | 0.13 | 0.06 | -0.04 | -0.04 | -0.02 | 0.21 | 1.00 | 0.06 | 0.04 | 0.03 | 0.02 | 0.09 | 0.06 | 0.04 | 0.03 |
| Diabetes | -0.16 | 0.00 | 0.02 | 0.07 | 0.07 | 0.03 | -0.00 | -0.21 | -0.16 | 0.04 | 0.06 | 1.00 | 0.12 | 0.09 | 0.04 | 0.13 | 0.06 | 0.14 | 0.08 |
| Heart Attack | -0.16 | 0.06 | 0.01 | 0.05 | 0.06 | 0.03 | 0.00 | -0.15 | -0.03 | 0.08 | 0.04 | 0.12 | 1.00 | 0.17 | 0.03 | 0.11 | 0.07 | 0.10 | 0.13 |
| Stroke | -0.13 | 0.00 | -0.01 | 0.05 | 0.06 | 0.02 | 0.01 | -0.14 | -0.02 | 0.05 | 0.03 | 0.09 | 0.17 | 1.00 | 0.04 | 0.10 | 0.06 | 0.08 | 0.10 |
| Asthma | 0.05 | -0.07 | -0.05 | 0.02 | 0.05 | 0.02 | 0.06 | -0.13 | -0.09 | 0.03 | 0.02 | 0.04 | 0.03 | 0.04 | 1.00 | 0.09 | 0.01 | 0.04 | 0.19 |
| Arthritis | -0.37 | -0.10 | 0.06 | 0.06 | 0.10 | 0.09 | -0.00 | -0.25 | -0.13 | 0.11 | 0.09 | 0.13 | 0.11 | 0.10 | 0.09 | 1.00 | 0.14 | 0.12 | 0.18 |
| Cancer | -0.24 | -0.03 | 0.05 | -0.02 | 0.02 | 0.06 | -0.04 | -0.11 | 0.00 | 0.05 | 0.06 | 0.06 | 0.07 | 0.06 | 0.01 | 0.14 | 1.00 | 0.09 | 0.08 |
| Kidney | -0.12 | -0.01 | 0.00 | 0.03 | 0.05 | 0.02 | 0.00 | -0.14 | -0.05 | 0.03 | 0.04 | 0.14 | 0.10 | 0.08 | 0.04 | 0.12 | 0.09 | 1.00 | 0.09 |
| COPD | -0.14 | -0.03 | -0.02 | 0.11 | 0.11 | 0.02 | 0.04 | -0.22 | -0.04 | 0.18 | 0.03 | 0.08 | 0.13 | 0.10 | 0.19 | 0.18 | 0.08 | 0.09 | 1.00 |

*Table S3: Comprehensive Interpretability vs. Performance Benchmark (Axis 2: BRFSS 2024 Temporal Cohort)*

| Algorithm | AUC | Brier Score | Algorithmic Transparency |
| --- | --- | --- | --- |
| MICE-EBM (Primary Proposed) | 0.8627 | 0.0845 | Exact / Intrinsic Glass-box |
| CatBoost | 0.8626 | 0.1399 | Post-hoc Only (SHAP) |
| LightGBM | 0.8624 | 0.1353 | Post-hoc Only (SHAP) |
| Stacked Meta-Ensemble (XGB+RF+LGBM) | 0.8622 | 0.0859 | Post-hoc Only (Black-box) |
| XGBoost | 0.8614 | 0.1348 | Post-hoc Only (SHAP) |
| MLP Deep Learning | 0.8604 | 0.0855 | Opaque (Black-box) |
| EBM + Median Imputation (Baseline) | 0.8594 | 0.0855 | Intrinsic Glass-box |
| Logistic Regression (Linear Benchmark) | 0.8572 | 0.1552 | High (Miscalibrated) |
| Random Forest | 0.8566 | 0.1382 | Post-hoc Only (SHAP) |

*Note: Models were trained on the BRFSS 2022 MICE-imputed derivation cohort and evaluated directly on the fully imputed BRFSS 2024 temporal cohort (n=414,644) to establish native distribution performance limits. No post-hoc recalibration was applied.*

***PART B: SUPPLEMENTARY FIGURES***

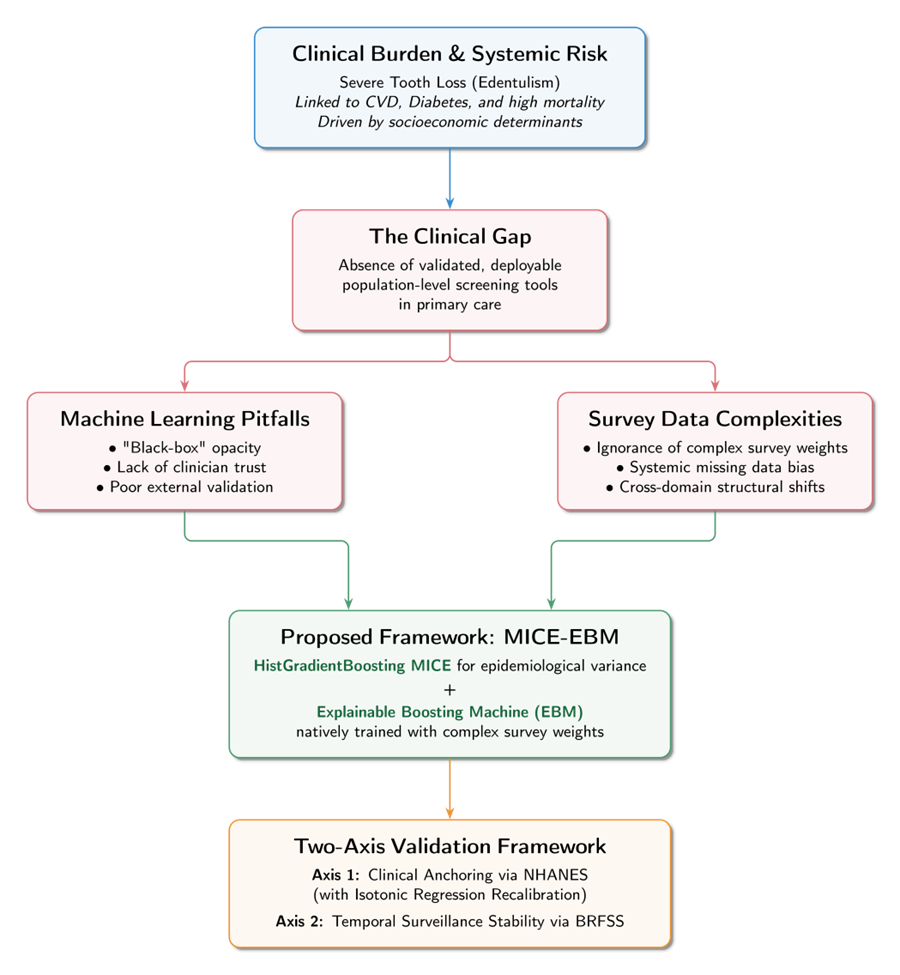

*Figure S1: Conceptual Framework and Motivation.*

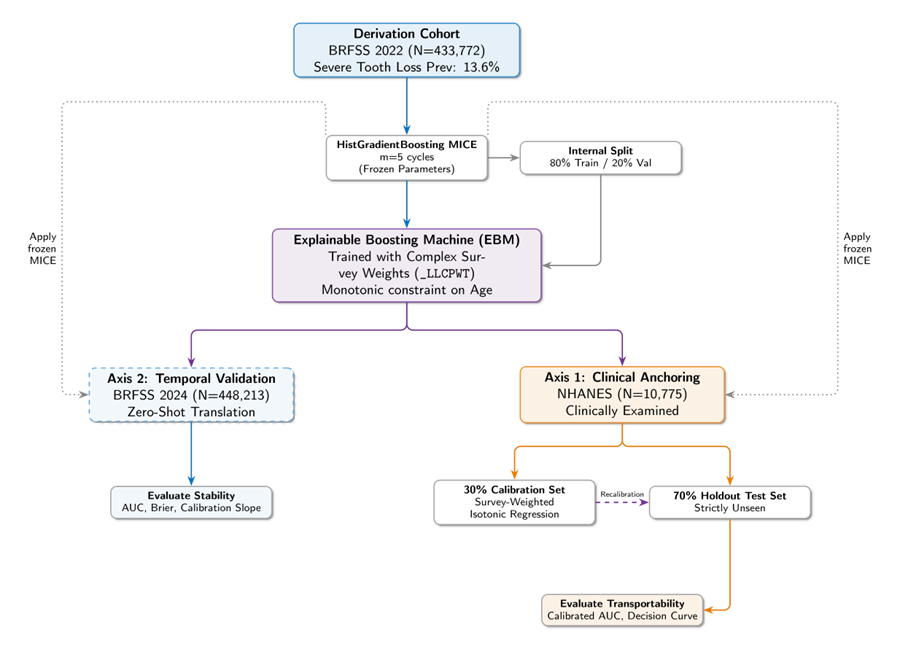

*Figure S2: Study Design and overall workflow.*

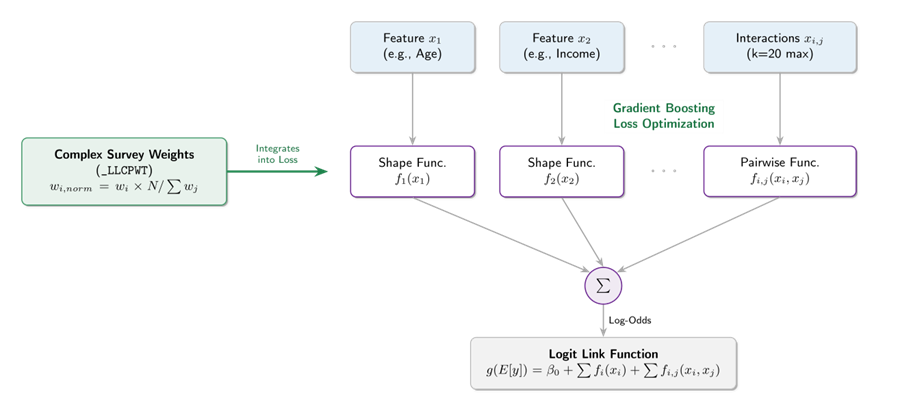

*Figure S3: Algorithm Architecture for the Explainable Boosting Machine.*

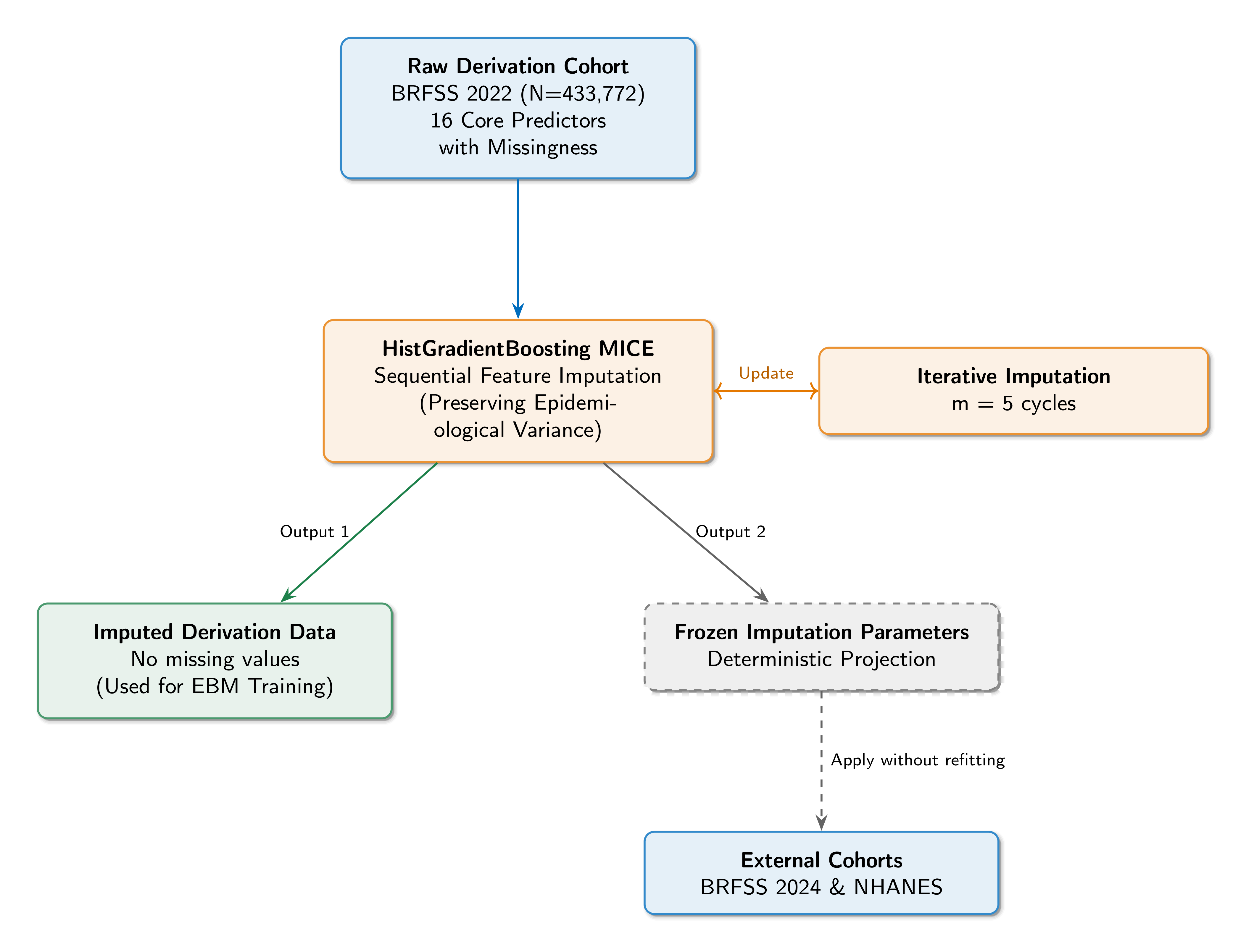

*Figure S4: HistGradientBoosting Multiple Imputation by Chained Equations (MICE) architecture.*

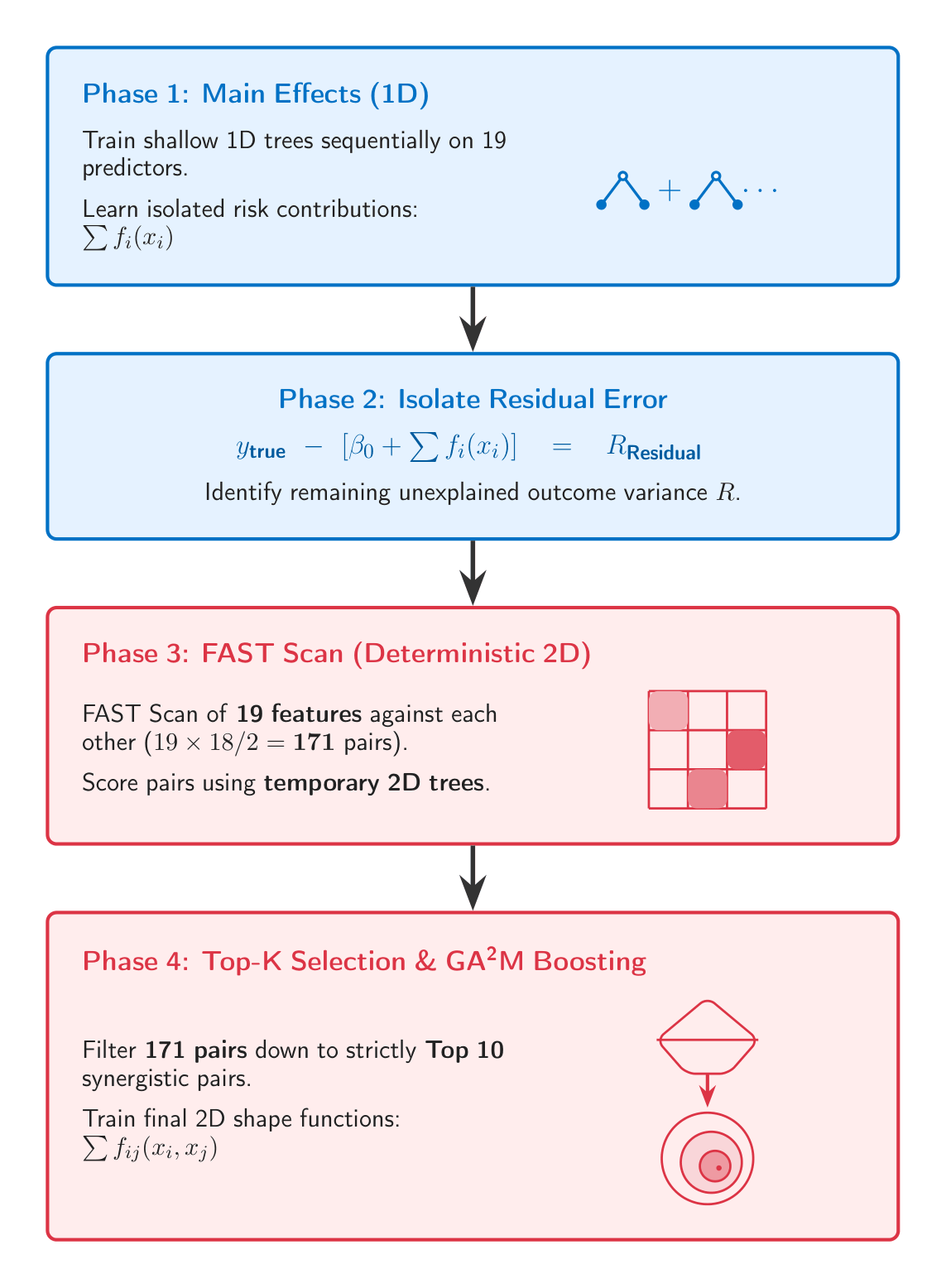

*Figure S5: FAST Interaction Pipeline*

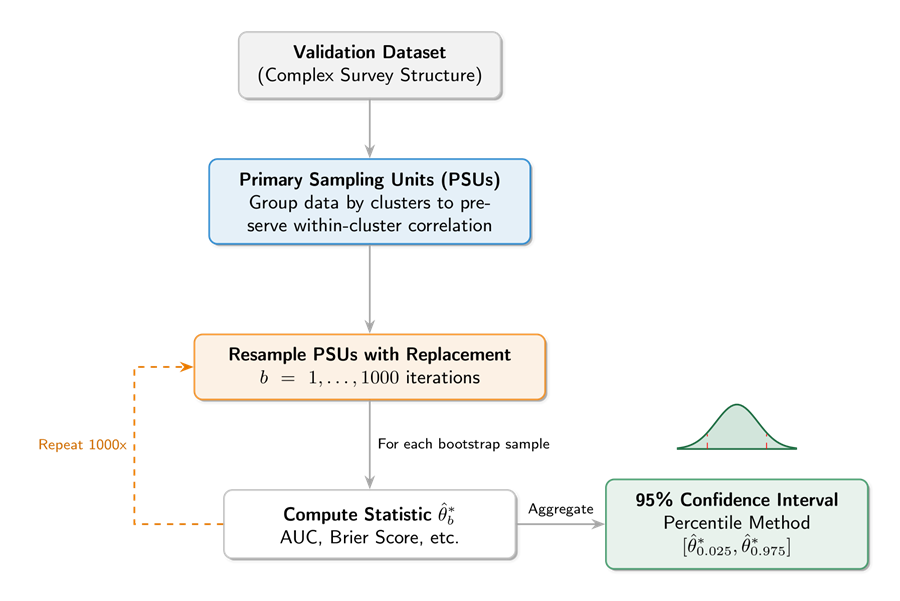
 *Figure S6: Stratified Cluster Bootstrapping process for rigorous interval estimation.*

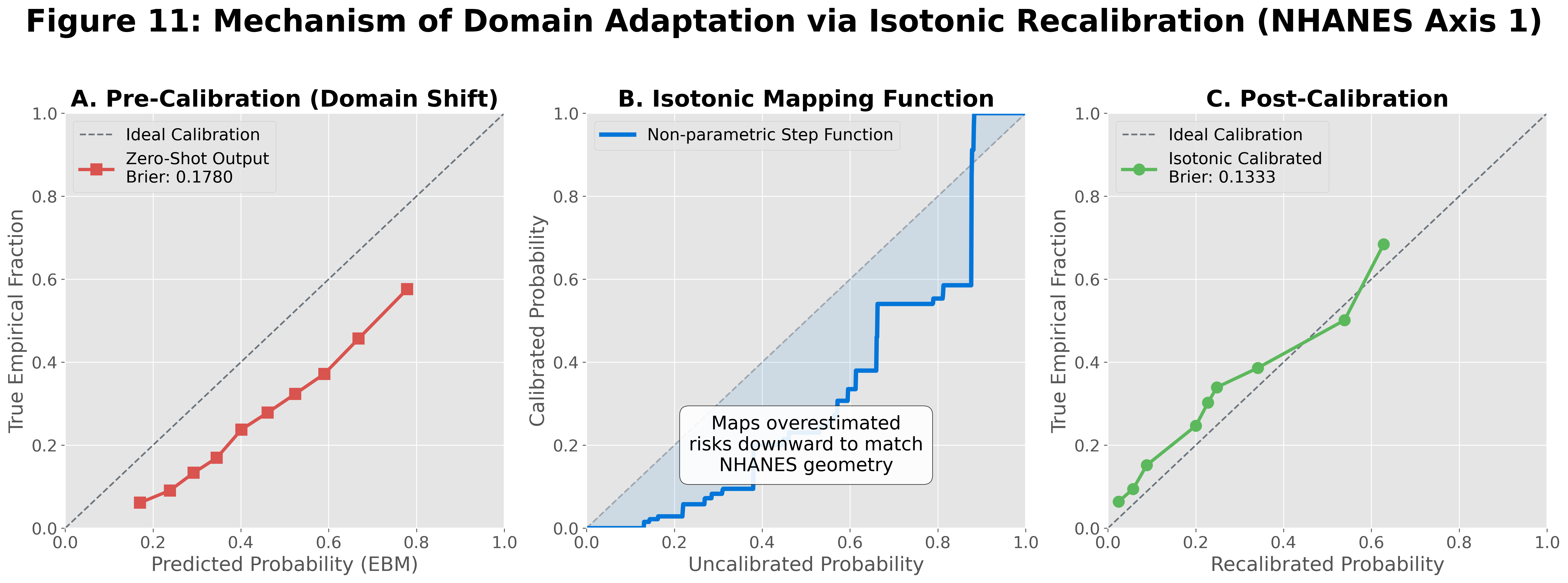

*Figure S7: Mechanism of Domain Adaptation via Isotonic Recalibration (Axis 1: NHANES Clinical Cohort).*

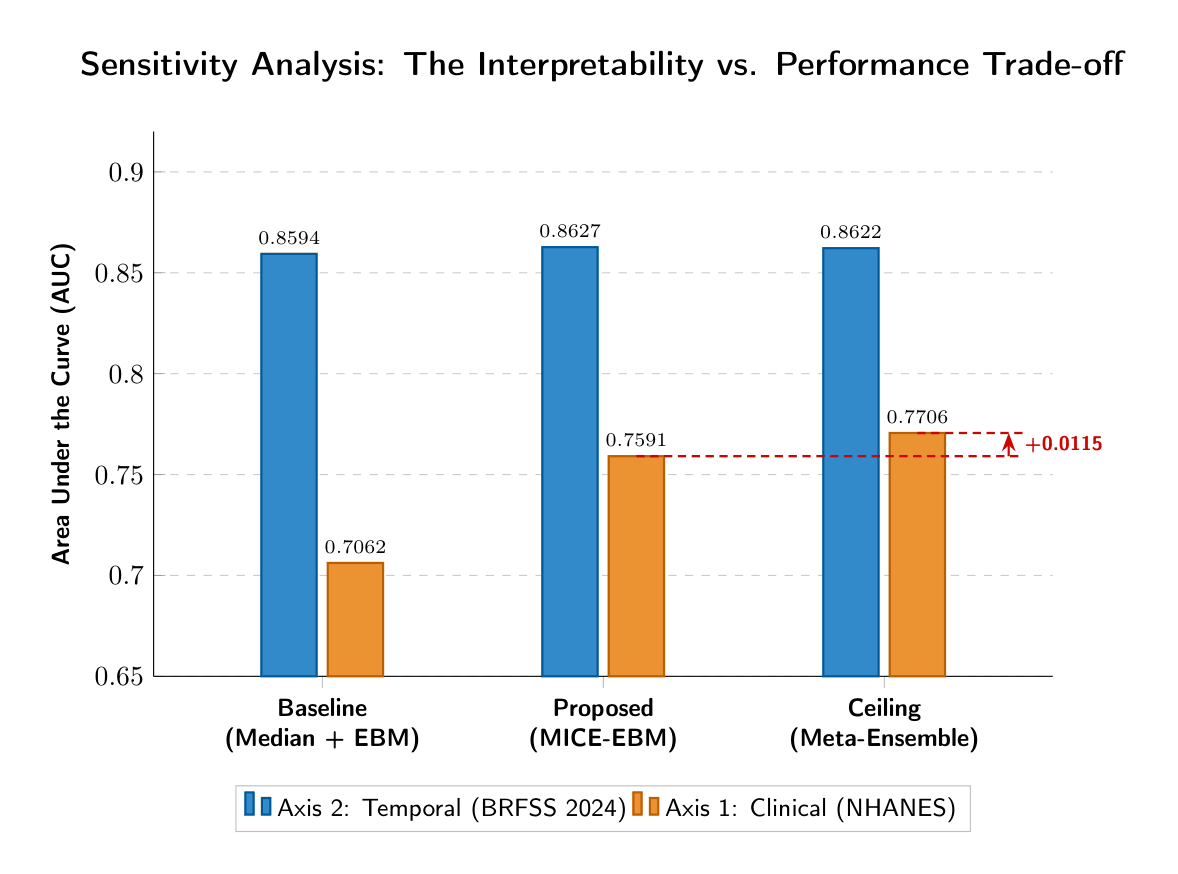

*Figure S8: Analysis of the tradeoff between raw predictive power and model explainability.*
